# Reduced circulating anti-CXCR3 antibodies as a common hallmark bridging systemic autoimmunity and atherosclerosis

**DOI:** 10.64898/2026.03.27.26349475

**Authors:** Daniel Miranda-Prieto, Mercedes Alperi-López, Ángel I. Pérez-Álvarez, Silvia Suárez-Díaz, Sara Alonso-Castro, Harald Heidecke, Ana Suárez, Gabriela Riemekasten, Javier Rodríguez-Carrio

**Affiliations:** Area of Immunology, Department of Functional Biology, Faculty of Medicine, University of Oviedo, Oviedo, Spain; Basic and Translational Research on Inflammatory Diseases, Department of Metabolism, Instituto de Investigación Sanitaria del Principado de Asturias (ISPA), Oviedo, Spain; Department of Rheumatology, Hospital Universitario Central de Asturias, Oviedo, Spain; Department of Neurology, Hospital Universitario Central de Asturias, Oviedo, Spain; Department of Internal Medicine, Hospital Universitario Central de Asturias, Oviedo, Spain; CellTrend GmbH, Luckenwalde, Germany; Department of Rheumatology and Clinical Immunology, University Medical Center Schleswig-Holstein Campus Lübeck, Lübeck, Germany

## Abstract

**Background/aims:** immune dysregulation underlies cardiovascular risk excess in systemic autoimmune diseases, such as rheumatoid arthritis (RA) and Sjögren disease (SjD). However, exact mediators are unknown. Regulatory autoantibodies targeting G protein–coupled receptors, including CXCR3, have emerged as modulators of immune and vascular homeostasis, but their role in autoimmunity remains ill-defined. Our aim was to evaluate anti-CXCR3 levels in systemic autoimmunity and their potential value as biomarkers.

**Methods:** anti-CXCR3 IgG serum levels were quantified in early RA (n=84), clinically-suspect arthralgia (n=12), and controls (n=65). Established RA (n=103) and SjD (n=44) were recruited for validation. Atherosclerosis was assessed by carotid ultrasound. Cytokines were measured by multiplex immunoassays. Cardiometabolic-related proteins were evaluated using high-throughput targeted proteomics. Publicly available datasets were used for validation.

**Results:** anti-CXCR3 antibodies were significantly reduced in early RA and arthralgia compared with controls, independently of disease activity, autoantibodies, or systemic inflammation. This finding was confirmed in validation cohorts. Anti-CXCR3 were negatively associated with good therapeutic outcomes upon csDMARD at 6 and 12 months. Lower anti-CXCR3 levels were independently associated with atherosclerosis occurrence and extent across conditions. Incorporating anti-CXCR3 into mSCORE improved risk stratification. Anti-CXCR3 were related to proteomic signatures linked to immune activation and to apoptosis, chemotaxis, and cell adhesion in an atherosclerosis-dependent manner. Transcriptomic analyses indicated compartment-specific CXCR3 dysregulation.

**Conclusion:** reduced anti-CXCR3 antibodies represent a shared hallmark bridging systemic autoimmunity and atherosclerosis burden, shaping our understanding on the regulatory role of antibodies at the vascular–immune interface. Clinical translation of anti-CXCR3 antibodies hold promise to improve risk stratification.

## INTRODUCTION

Autoimmune diseases, such as rheumatoid arthritis (RA) or Sjögren disease (SjD) are hallmarked by a substantial cardiovascular disease (CVD) risk excess [1]. Chronic immune dysregulation and autoimmune phenomena are thought to underlie this risk excess, and accumulating evidence points to shared mechanisms among conditions [1,2]. However, precise immunopathogenic pathways linking systemic autoimmunity and atherosclerosis remain only partially understood.

Humoral immune responses have been proposed as central contributors to CVD [3]. Increased levels of autoantibodies found in atherosclerosis mice model suggests an active role as drivers of atherosclerosis, but antigen-antibody axes are ill-defined [1,4], and B-cell subset modulation led to opposed, even contradictory results, hence adding another layer of complexity of antibody-mediated regulation in vascular disease.

Autoantibodies against G-coupled protein receptors (GPCRs) have recently emerged as central players of immune and tissue homeostasis and key contributors to different pathologic conditions such as autoimmune diseases, infections, and neuroimmune disorders [5]. These autoantibodies can modulate GPCRs signalling through activation, inhibition or biased activation of GPCRs or their ligands [6], thereby fine-tuning receptor activity and downstream cellular responses. Integrative studies using omic approaches may help capturing functional relevance of these antibodies.

GPCRs are involved in a broad range of biological processes related to atherosclerosis. Autoantibodies against GPCRs such as angiotensin II type-1 receptor, involved in vascular damage and systemic inflammation, and against β1AR, a GPCR expressed in cardiac and immune cells, have been associated with CVD (reviewed in [5,7]). More recently, antibodies targeting CXCR3 have been shown to predict CV risk [8], likely through skewed Th1 activation. Although already linked to lung manifestations in systemic sclerosis (SSc) [9], evidence on the role of anti-CXCR3 in systemic autoimmune diseases is rather limited, especially in RA. Moreover, whether altered levels of these antibodies can be found in the earliest stages of the disease remains unexplored.

Given the unmet need for biomarkers that integrate systemic immune dysregulation and atherosclerosis in systemic autoimmunity, anti-CXCR3 antibodies represent a promising candidate. Therefore, the main aims of the present study were to assess anti-CXCR3 antibody levels in RA and SjD patients, to evaluate potential associations with atherosclerosis burden, and to investigate their role as a biomarker for CV risk stratification.

## MATERIAL AND METHODS

### Study participants

Early RA patients fulfilling 2010 CR/EULAR classification criteria [10] were recruited at disease onset from the early arthritic clinic of the Department of Rheumatology at Hospital Universitario Central de Asturias (HUCA). RA patients were not exposed to any disease-modifying antirheumatic drugs (DMARDs) at recruitment. A complete clinical examination was performed during the clinical appointment, including Disease Activity Score 28-joints and Health Assessment Questionnaire (HAQ) calculations. Traditional CV risk factors (dyslipidaemia, hypertension, diabetes, smoking habits and obesity) were obtained from the medical records and defined in compliance with national guidelines. A group of individuals with clinically suspect arthralgia (CSA) [11] were also recruited from the same clinic. A separate cohort of long-lasting, established RA patients recruited from the same institution was included as validation cohort [12]. Additionally, a group of 13 biological-naïve RA patients was prospectively followed upon TNF blockade initiation and a blood sample was obtained before (baseline) and 3 months after anti-TNF therapy. Clinical response after TNF blockade was evaluated according to EULAR criteria [13]. Healthy controls (HC) were recruited among age- and sex- matched healthy volunteers from the same population.

Furthermore, a cohort of SjD patients fulfilling American-European Consensus Group criteria [14] was recruited from the Department of Internal Medicine (HUCA). These patients undergone a complete clinical examination, including Sjögren’s syndrome disease activity index (ESSDAI) calculation, and traditional CV risk assessment as previously mentioned.

Fasting blood samples were collected from all individuals by venepuncture in EDTA-containing tubes. Blood samples were immediately transferred to the laboratory and processed within less than 2 hours. Serum samples were stored at −80°C until experimental procedures. Conventional blood biochemical, including C-reactive Protein (CRP) and Erythrocyte Sedimentation Rate (ESR) measurements, lipid analyses and complete blood counts were carried out in all individuals.

The study was approved by the local institutional review board (Comité de Ética de Investigación con Medicamentos del Principado de Asturias) in compliance with the Declaration of Helsinki (reference CEImPA 2021.126). All study subjects gave written informed consent.

### Echocardiographic study

Subclinical atherosclerosis was assessed by Doppler ultrasound at the sonography laboratory (Department of Neurology, HUCA) online in B-mode using a Toshiba Aplio XG device (Toshiba American Medical Systems) by an experienced user, as previously described [15]. The carotid intima-media thickness (cIMT) was bilaterally measured following the “Mannheim Carotid Intima-Media Thickness Consensus 5 (2004-2006) [16], and atherosclerotic plaque burden (presence and extent) were defined according to established definitions as previously described [16]. Plaque vulnerability status was evaluated by the ultrasound characterization of the plaques (border profile, echo-density, calcification, and burden), and these were classified as “low” or “high” risk [17].

### Anti-CXCR3 assessment

Levels of anti-CXCR3 IgG antibodies were quantified in serum samples (diluted 1/100) using a commercial assay (EIA for Quantitative Determination of anti-CXCR3-Antibodies, CellTrend), following the instructions provided by the manufacturer. Values were expressed as U/ml, according to standard curves. Detection limit was 2.5 U/ml. Intra- and inter-assay coefficient of variation were 6.1% and 8.0%, respectively.

### Cytokine analyses

Interleukin (IL)-6, Tumour Necrosis Factor (TNF), Interferon gamma (IFNγ), IL-1b, IL-23, IL-12, IL-33, IL-10, IL-17, IL-8, and IL-18 serum levels were measured by a predefined multiplex assay (Human Inflammation Panel 1, LEGENDplex, BioLegend). Detection limits were 2.68 pg/ml, 2.90 pg/ml, 2.68 pg/ml, 3.17 pg/ml, 4.39 pg/ml, 19.53 pg/ml, 3.66 pg/ml, 0.73 pg/ml, 2.92 pg/ml, and 3.90 pg/ml, respectively. The serum levels of IFNα were quantified using a Cytometric Bead Array Flex Set (BD). Detection limit was 1.25 pg/ml, A Proliferation -Inducing Ligand (APRIL), B-cell Activating Factor (BAFF) and IL-21 serum levels were quantified using ELISA kits (Invitrogen, ThermoFisher). Detection limits were 0.40 ng/ml, 0.13 ng/ml and 20 pg/ml, respectively.

### Proteomic platform

Serum proteomics were carried out using a high-throughput analysis by means of the Proximity Extension Assay [18]. In brief, a predefined panel of 92 protein species related to CV was measured using Olink technology (Cardiovascular Panel II) (Olink Bioscience, Sweden).

### Statistical analysis

Variables were summarised as median (interquartile range) or mean ± standard deviation, or *n* (%), as appropriate.

Differences among groups were evaluated by Mann-Whitney *U*, Kruskal–Wallis tests with Dunn-Bonferroni correction for multiple comparisons, or Wilcoxon paired tests, according to the distribution of the variables. Correlations were assessed by Spearman ranks’ test. The associations between anti-CXCR3 levels and atherosclerosis occurrence were analyzed by univariate and multivariate logistic regression adjusted by confounders. Negative binomial regression models were used when plaque number was modeled as outcome variable. Odds ratio (OR) and 95% confidence intervals (CI) were calculated. Anti-CXCR3 IgG levels were added to the mSCORE by using tertiles observed in the HC population as cut-offs, with scorings been given as follows [19]: 0 to T1: +2, T1 to T2: +1, T2 to T3: +0. These points were added to the mSCORE values to generate the modified algorithm (mSCORE + anti-CXCR3). The performance on risk stratification was evaluated by classification (sensitivity, specificity, accuracy), discrimination (AUC ROC), correlation (Matthews’ correlation coefficient), and goodness of fit (Hosmer-Lemeshow test) statistics. Differences between ROC values were compared by De Long test. Net Reclassification Improvement (NRI) was used to compare the effect of risk reclassification of the modified algorithm.

Protein-protein interactions were assessed by the STRING database [20], and pathway enrichment analyses were evaluated by pathway using Metascape [21] using KEGG pathway, GO biological processes, Reactome Gene Sets, Canonical Pathways, CORUM, and PANTHER Pathway as ontology sources. Term network analyses were performed under Cytoscape [22]. Group-wise comparisons were performed under Metascape and visualized using circus plots, and heatmap of enriched terms with hierarchical clustering.

Gene expression datasets were downloaded from the publicly available National Center for Biotechnology Information (NCBI) Gene Expression Omnibus (GEO) repository [23]. For validation, differential CXCR3 expression was evaluated using the GEO2R tool and FDR-adjusted p-values (p(adj)) were computed. Then, profile data expression was downloaded, normalized (Z-score), and analyzed by conventional tests across groups.

A p-value <0.050 was considered as statistically significant. Statistical analyses were carried out under SPSS v. 27, R v.4.1.3 and GraphPad Prism 8.0.

## RESULTS

### 1. Anti-CXCR3 antibodies in patients with systemic autoimmune diseases

A total of 82 early RA patients, 14 CSA individuals and 65 HC were recruited for this study (Supplementary Table 1). The levels of anti-CXCR3 antibodies were found to be decreased in early RA and CSA groups compared to HC (Figure 1A). This result was confirmed in the validation cohort (Figure 1B), composed by patients with established RA (Supplementary Table 2). Furthermore, equivalent findings were observed in the analysis of the SjD cohort (Figure 1C) (Supplementary Table 3).

**Figure 1:**
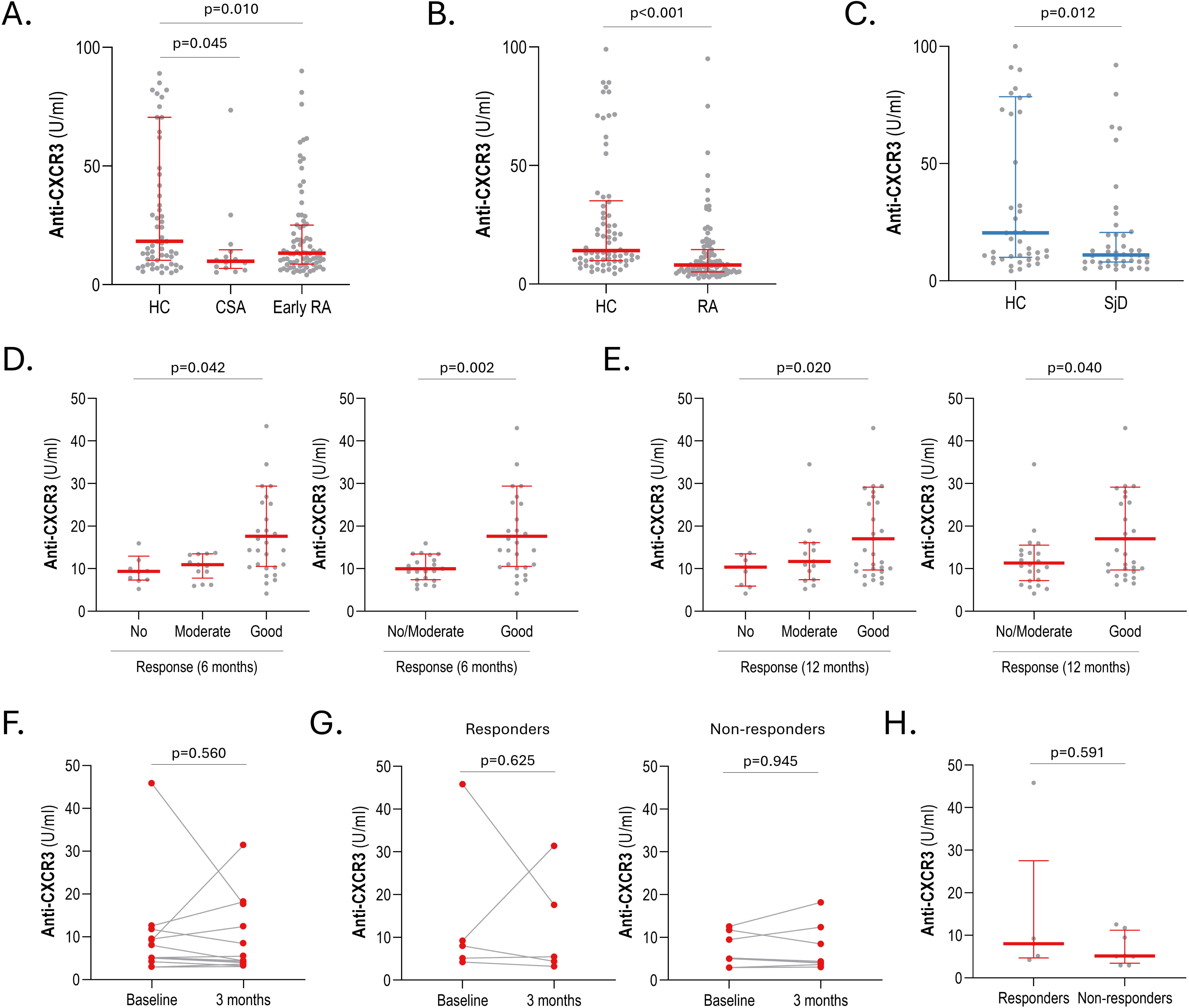
Anti-CXCR3 antibodies in systemic autoimmune diseases. The serum levels of anti-CXCR3 antibodies were compared in early RA, CSA and HC (A), established RA and HC (B), SjD patients and HC (C). The serum levels of anti-CXCR3 antibodies were compared in early RA patients according to clinical outcomes (EULAR criteria) at 6 (D) or 12 months (E) upon csDMARD inhibition. Anti-CXCR3 antibodies were measured at baseline and 3-months after TNF blockade initiation in biological-naïve RA patients (F), and according to clinical outcomes (G). Differences at baseline were also assessed (H). In scatter plots, each dot represents one individual. Bars represent 25th percentile (lower), median and 75th percentile (upper). Colour code depicts RA (red) or SjD (blue) data. Differences were assessed by Kruskal-Wallis (with Dunn-Bonferroni correction for multiple comparisons), Mann-Withney U tests, or paired Wilcoxon paired test, as appropriate. P-values were indicated.

Anti-CXCR3 antibodies were found to be unrelated to demographic and clinical features in early RA patients, including disease activity and symptom duration (Table 1). Disease-related autoantibodies (RF and ACPA) failed to show an association with anti-CXCR3 antibodies, whereas ANA positivity was associated with lower anti-CXCR3 levels (Table 1). No associations with the total level of immunoglobulins or complement levels were found (Table 1). Finally, anti-CXCR3 serum levels failed to show an association with proinflammatory cytokines, including B-cell related factors, in early RA (Supplementary Table 4).

**Table 1:**
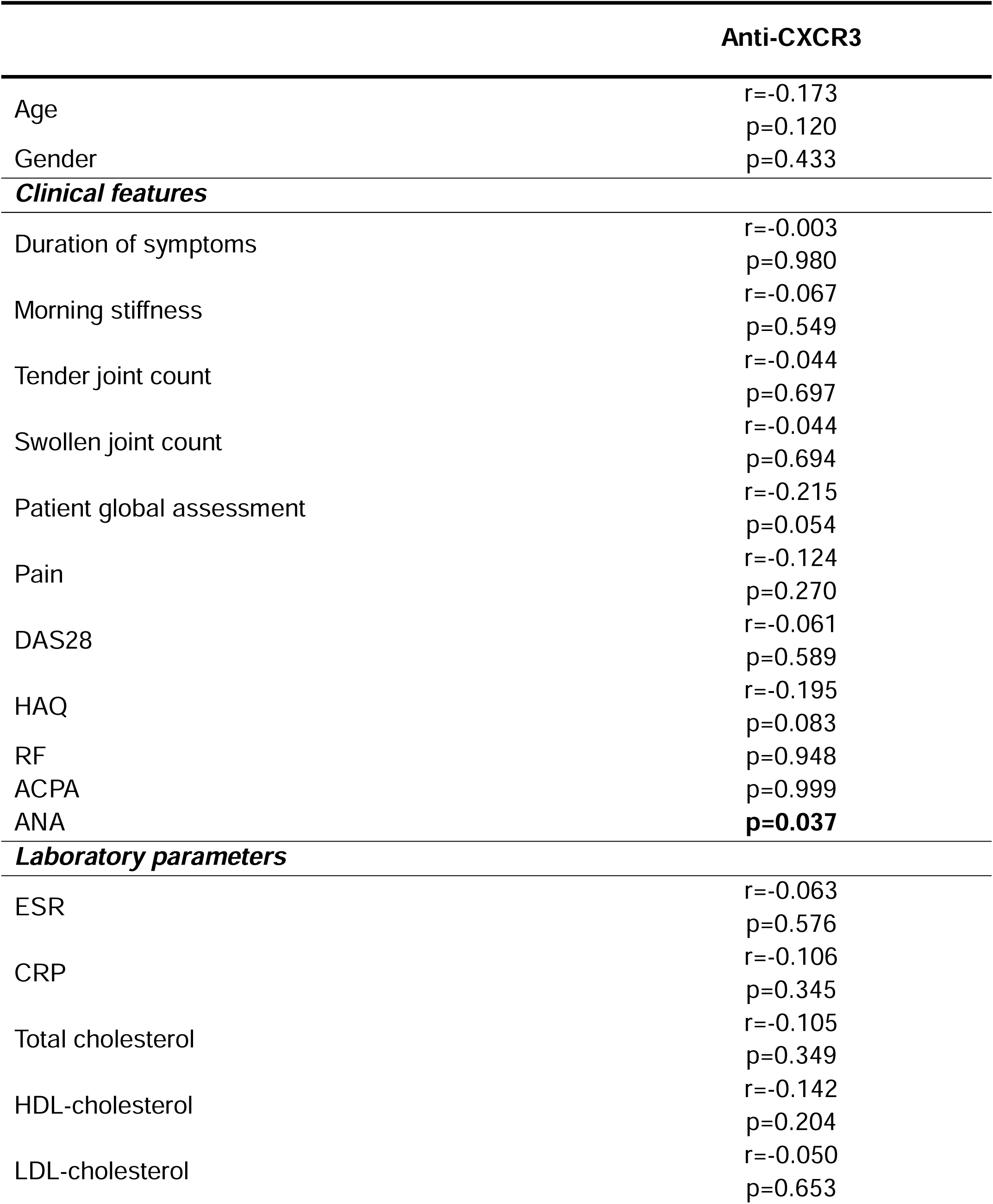
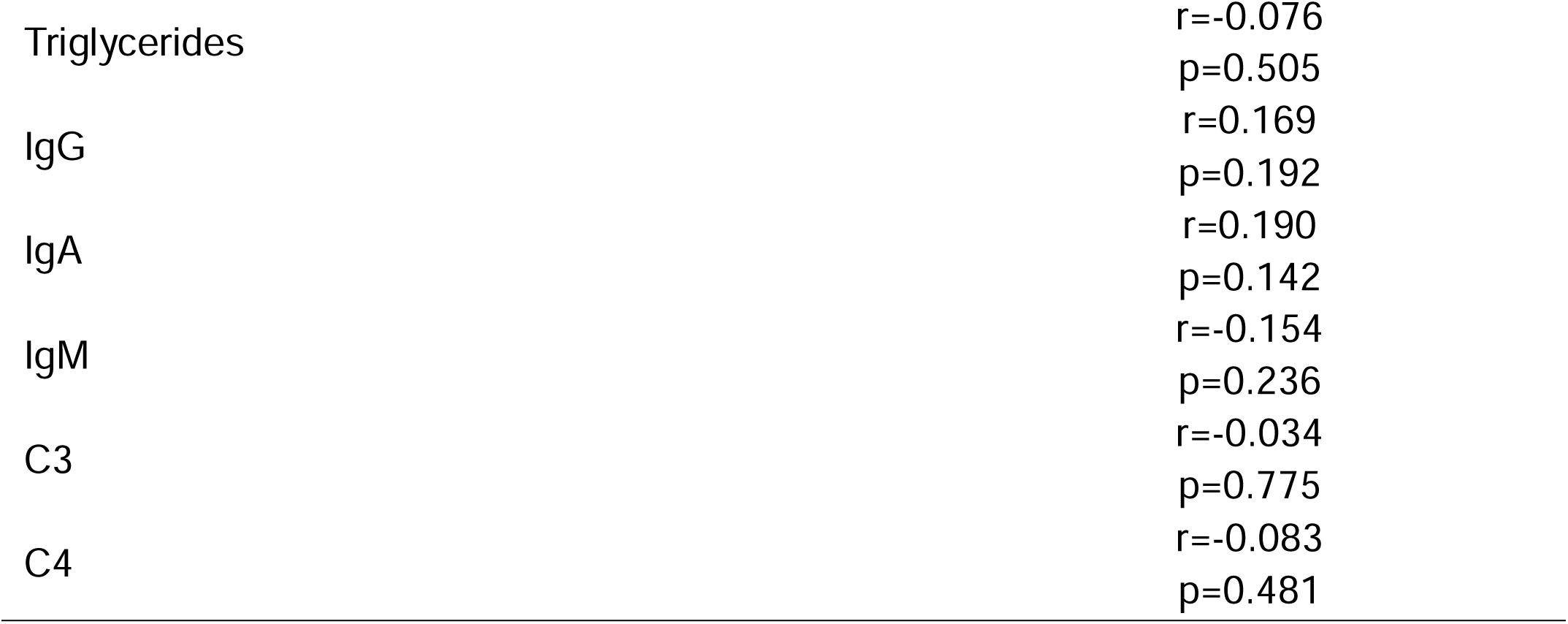
Anti-CXCR3 antibodies and clinical features in early RA. Associations between serum levels of anti-CXCR3 antibodies and demographic, clinical features, and laboratory parameters were analysed by Spearman’s rank tests or Mann-Withney U tests in early RA patients. Coefficients (r) and p-values, or p-values for the difference between groups are shown. Those reaching statistical significance were highlighted in bold.

Lower levels of anti-CXCR3 antibodies at baseline were observed in early RA patients experiencing poor therapeutic response at 6 months upon csDMARD initiation (n=28) compared to those exhibiting good responses (n=30) (Figure 1D). Equivalent results were observed at 12 months (Figure 1E). On the contrary, anti-CXCR3 levels were unchanged at 3 months upon TNFi initiation (Figure 1F) (Supplementary Table 5), irrespectively of treatment outcomes (Figure 1G-H).

Taken together, these findings suggest that reduced anti-CXCR3 levels may be a common hallmark across systemic autoimmune conditions. Decreased anti-CXCR3 antibodies were found along the whole RA continuum, including the earliest stages, and connected with therapeutic outcomes.

### 2. Anti-CXCR3 antibodies and atherosclerosis burden in autoimmunity

Next, whether circulating anti-CXCR3 levels were associated with atherosclerosis burden and risk factors was investigated.

Anti-CXCR3 levels did not exhibit associations with traditional CV risk factors (hypertension: p=0.085, dyslipidemia: p=0.345, diabetes: p=0.876, smoking: p=0.075, and obesity: p=0.789) in early RA. Patients with atherosclerosis exhibited lower anti-CXCR3 antibodies than their atherosclerosis-free counterparts (Figure 2A). Patients with high-risk plaques according to their echocardiographic characteristics exhibited lower anti-CXCR3 levels than those presenting with low-risk plaques (15.19 (13.28) vs 10.24 (7.09) U/ml, p=0.002). Furthermore, anti-CXCR3 levels were negatively associated with plaque number (r=0.248, p=0.020), although no associations were found with cIMT (r=0.139, p=322). The analysis of CVD history in the validation cohort confirmed these findings (Figure 2B). Regression analyses demonstrated that anti-CXCR3 levels were independent predictors of atherosclerosis occurrence in univariate or multivariate models in early RA, after adjusting for traditional CV risk factors (Table 2). Equivalent results were obtained when plaque number was modeled in fully-adjusted multivariate negative binomial regression analyses (b [95% CI]: −0.026 [−0.032, −0.010], p=0.045), thereby confirming the independent association between anti-CXCR3 levels and atherosclerosis presence and extent in early RA. Moreover, a similar picture was also obtained in SjD (Figure 2C) (Supplementary Table 6).

**Figure 2:**
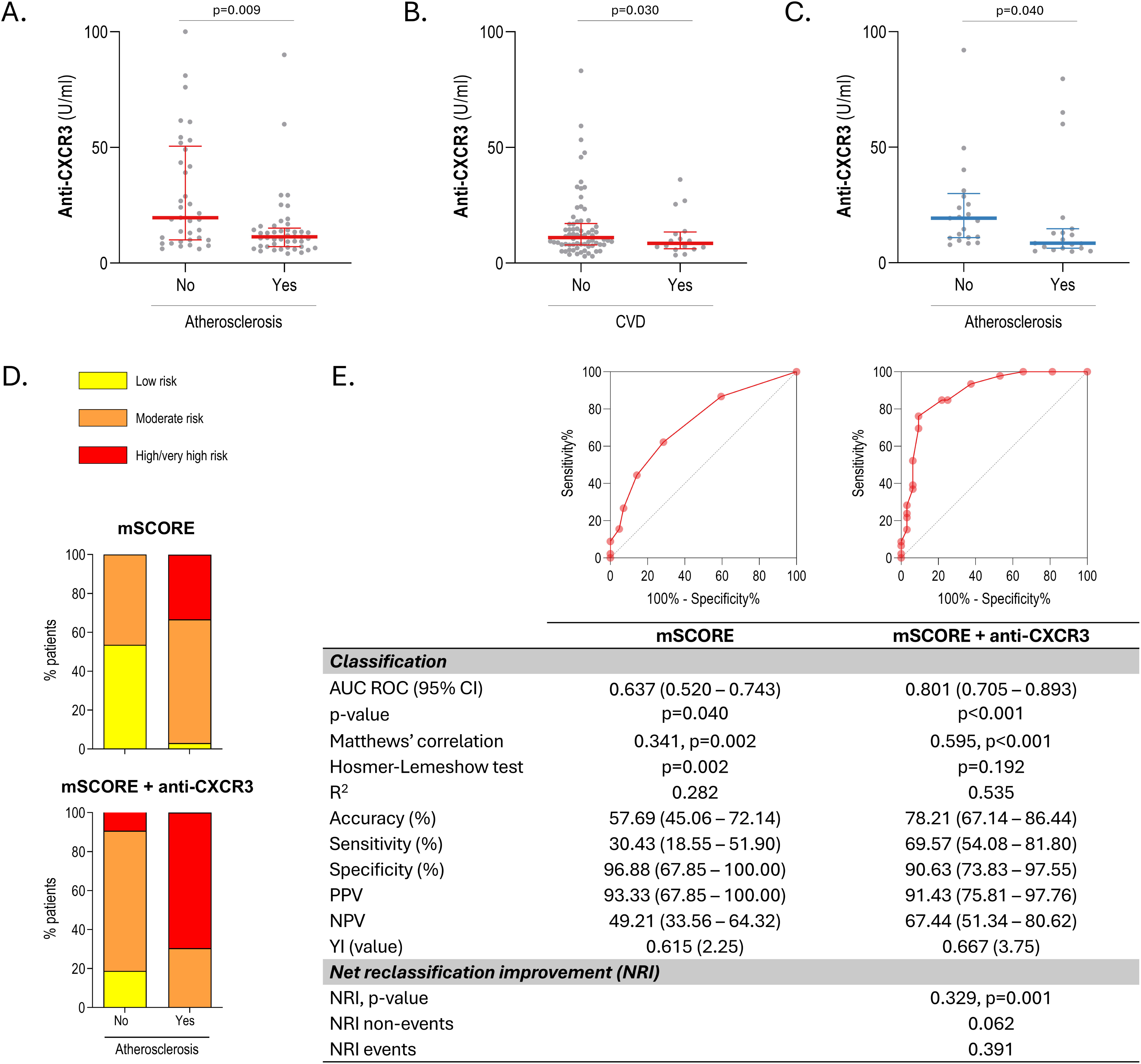
Anti-CXCR3 and atherosclerosis burden in systemic autoimmune diseases. The serum levels of anti-CXCR3 antibodies were compared according to atherosclerosis status in early RA patients (A), according to CVD status in established RA (B), and atherosclerosis status in SjD (C). Bars represent 25th percentile (lower), median and 75th percentile (upper). Colour code depicts RA (red) or SjD (blue) data. Differences were assessed by Mann-Withney U tests. P-values were indicated. (D) Adding anti-CXCR3 strata to existing instrument (mSCORE + anti-CXCR3) improved risk stratification in early RA into more realistic risk groups, particularly in patients with atherosclerosis, compared to original tool (mSCORE), based on risk category distribution (low, moderate, and high/very high) and atherosclerosis status. (E) Classification, diagnostic metrics and reclassification measures (NRI) were calculated to evaluate incremental value. ROC curves for each model are shown at the top of the table.

**Table 2:** Anti-CXCR3 antibodies as predictors of atherosclerosis in RA. The role of anti-CXCR3 levels as predictor of atherosclerosis occurrence in early RA patients was analysed by univariate and multivariate logistic regression models. Odds ratio (OR), 95% confidence intervals (95% CI) and p-values for each predictor were shown. Those reaching statistical significance were highlighted in bold.

|  | OR | 95% CI | p-value |
| --- | --- | --- | --- |
| ***Univariate*** |  |  |  |
| Anti-CXCR3, per unit | 0.967 | 0.942 – 0.993 | **0.012** |
| ***Multivariate*** |  |  |  |
| Anti-CXCR3, per unit | 0.939 | 0.890 – 0.990 | **0.019** |
| Sex, women | 0.219 | 0.034 – 1.412 | 0.111 |
| Age, per year | 1.081 | 1.010 – 1.157 | **0.025** |
| Smoking, yes | 3.728 | 0.924 – 15.030 | 0.064 |
| Hypertension, yes | 1.751 | 0.395 – 7.778 | 0.461 |
| Diabetes, yes | 0.001 | 0.000 – 0.001 | 0.998 |
| Dyslipidemia, yes | 1.395 | 0.924 – 5.973 | 0.654 |

Our analyses revealed that anti-CXCR3 alone were able to discriminate between early RA patients with and without atherosclerosis (AUC ROC [95% CI], p: 0.671 [0.545-0.797], p=0.009). We therefore evaluated if anti-CXCR3 antibodies could be useful in risk stratification. Adding anti-CXCR3 tertiles to the mSCORE (mSCORE + anti-CXCR3), improved risk stratification into more realistic risk categories (Figure 2D). The analysis of diagnostic and classification statistics supported this observation (Figure 3B). A better discrimination capacity (difference between areas = 0.164 [0.008–0.247], p<0.001), and improved classification metrics (accuracy, sensitivity, and Matthews’ Correlation coefficient) (Figure 2E) was achieved. NRI analysis confirmed a better reclassification performance, with a noticeable effect for those presenting atherosclerosis and a marginal to no effect in the atherosclerosis-free group (Figure 2E).

**Figure 3:**
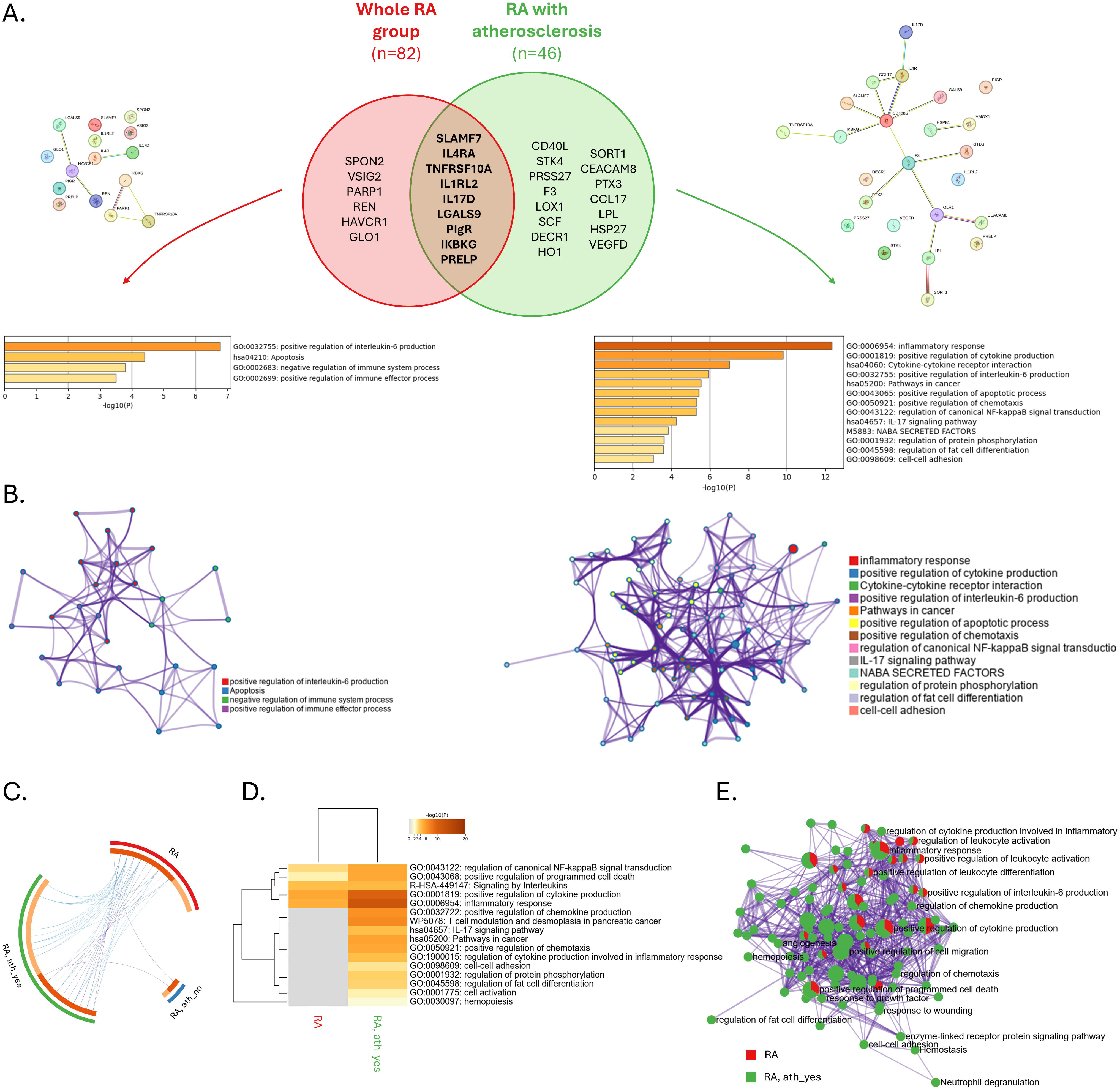
Proteomic analyses of anti-CXCR3 autoantibodies in RA and atherosclerosis status. Protein hits found to be associated with anti-CXCR3 antibody serum levels in the whole RA group and in the subgroup of RA patients with atherosclerosis were analyzed. Overlap analysis was shown as a Venn diagram (A). Shared and group-specific proteins are listed in the overlap and non-overlap regions, respectively. Protein-protein interaction networks for the corresponding protein sets were generated using STRING. Pathway and process enrichment analysis was conducted under Metascape using KEGG pathway, GO biological processes, Reactome Gene Sets, Canonical Pathways, CORUM, and PANTHER Pathway as ontology sources. P-values for each term was calculated based on the accumulative hypergeometric distribution. To further capture the relationships among terms, enriched terms were selected and rendered as network plots (B), where terms with similarity>0.3 were connected by edges. The networks were visualized using Cytoscape, where each node represented an enriched term and it is depicted by its cluster ID (see colour legend). For group-wise analyses, circus plots (C) were used to shown to visualize and compare protein overlaps among the whole RA group, and patient subgroups according to atherosclerosis status (outside arcs). Inner arcs depicted protein usage: shared (dark orange) and unique (light orange). Purple lines link proteins shared by multiple groups. The greater the number of purple links and the longer the dark orange arcs implies greater overlap among groups. Blue lines link proteins that, although different, fall under the same ontology term across groups (that is, functional overlap among protein sets). To compare pathway and process enrichment patterns among groups, a heatmap of enriched terms with hierarchical clustering (kappa threshold: 0.3) was produced (D). The heatmap cells were coloured by their p-values. Integrated network layout of enriched terms across groups was derived to highlight shared and group-specific functional modules (E). Each term is represented by a node, where size is proportional to the number of protein sets that fall under that term, and colour code represents a pie chart proportional to number of hits from the protein sets. Main terms were labelled. Colour code depicts whole RA group (red) or RA patients with atherosclerosis (green) and RA patients without atherosclerosis (green) data. RA, whole RA group; ath_yes, RA patients with atherosclerosis; ath_no, RA patients without atherosclerosis.

All these findings reinforce an independent association between decreased anti-CXCR3 antibodies and atherosclerosis in autoimmune conditions, also exhibiting an incremental value for risk stratification over existing instruments.

### 3. Anti-CXCR3 antibodies and proteomic signatures

Next, in order to get a more functional insight into the mechanisms behind anti-CXCR3 alterations, the associations between anti-CXCR3 levels and serum proteomic signatures were evaluated.

The levels of anti-CXCR3 antibodies were correlated with a number of protein hits in early RA (n=15). These proteins exhibited a significant protein-protein interaction (p=0.005, local average clustering coefficient=0.467) (Figure 3A, left). Pathway process enrichment analysis using Metascape uncovered a total of four functional pathways participated by these proteins, overall related to immune activation processes (Figure 3A). Subgroup analyses according to atherosclerosis status revealed a larger set of proteomic correlates (n=24) in patients with atherosclerosis, which were also strongly interconnected (PPI: p=1.25·10-5, local average clustering coefficient=0.475) (Figure 3A, right). Proteomic correlates in the atherosclerosis-free group were negligible (n=3). Protein traits differentially associated with the atherosclerosis group included species functionally related with CVD (LOX-1, F3, HO-1, SPRT1, CEACAM8, PTX3, LPL, HSP27 and VEGFD), in addition to those related to broad immunological processes. Pathway analyses restricted to this patient subset identified a number of events intimately linked to atherosclerosis, such as apoptosis, chemotaxis, fat cell differentiation and cell adhesion, as well as processes related to intracellular signalling pathways (Figure 3A, right).

Network analyses of terms retrieved in pathway and process enrichment analysis highlight clear differences in the structure and coherence of the biological signals (Figure 3B). The network derived from the whole RA group showed fewer connections and more dispersed functional terms, suggesting a broader and less coordinated set of processes. In contrast, the network derived from the group with atherosclerosis displayed a larger number of enriched processes with stronger interconnectivity, forming tighter clusters of related biological functions. This increased density reflects greater functional convergence among the top-ranked terms and more consistent enrichment signals across clusters, thereby indicating a more robust and biologically coherent pathway landscape in the latter.

Group-wise comparisons strengthened these findings. Of note, a significant overlap was confirmed, thus indicating that patients with atherosclerosis mostly recapitulated the proteomic set observed at the whole RA group level (Figure 3C), whereas the effect of atherosclerosis-free group was marginal. Pathways and process analyses revealed two main clusters, one being common to whole RA group and RA with atherosclerosis group (broad immune activation and inflammation), whereas the second one was specific to the patient subset with atherosclerosis and included functionally-related signatures (Figure 3D). Network analysis supported this differential usage of retrieved terms between groups (Figure 3E).

The analysis of the SjD cohort yielded equivalent findings compared to RA. Although proteomic hits associated with anti-CXCR3 in SjD (n=10) (Supplementary Figure 1A) slightly diverged from those in RA, pathways retrieved were overall similar (Supplementary Figure 1B). Group-wise analyses confirmed overlap in pathway and process enrichment analyses (Supplementary Figure 1C-D). Interestingly, most of the proteomic correlates were overall recapitulated (6/10) by the group of patients with atherosclerosis in SjD (n=22) (Supplementary Figure 1E).

Altogether, these findings suggest that anti-CXCR3 antibodies were associated with proteomic signatures linked to immune activation and to apoptosis, chemotaxis, and cell adhesion in an atherosclerosis-dependent manner. This effect was consistent across systemic autoimmune conditions.

### 4. CXCR3 expression at systemic and tissue level in systemic autoimmunity

Next, in order to get insight into anti-CXCR3 alterations, the expression of CXCR3 in systemic autoimmunity was evaluated.

Data from CXCR3 gene expression both in the systemic compartment was well as in target tissue (synovium) was extracted from publicly available microarray datasets from the GEO database. Four datasets containing relevant samples for synovial tissue analysis were found. GSE1919 included gene expression data from 5 RA patients, 5 OA patients and 5 controls. A differential gene expression for CXCR3 was obtained (p(adj)=0.023), thus demonstrating an enhanced CXCR3 expression in the RA group (Figure 4A). Analysis of the datasets GSE55457 (13 RA, 10 OA, 10 controls) (Figure 4B) and GSE55235 (13 RA, 10 OA, 10 controls) (Figure 4B) confirmed these findings (p(adj)=0.010 and p(adj)=6.65·10-5, respectively). Differences between RA (n=10) and OA (n=6) were also replicated in the GSE55584 dataset (p=0.044) (Figure 4D). GSE36700 also confirmed the differential CXCR3 expression at the synovial level (p(adj)=0.011) (Figure 4E) compared to a broader range of disease groups (7 RA, 5 OA, 4 SLE, 4 microcrystalline arthritis, 4 seronegative arthritis).

**Figure 4:**
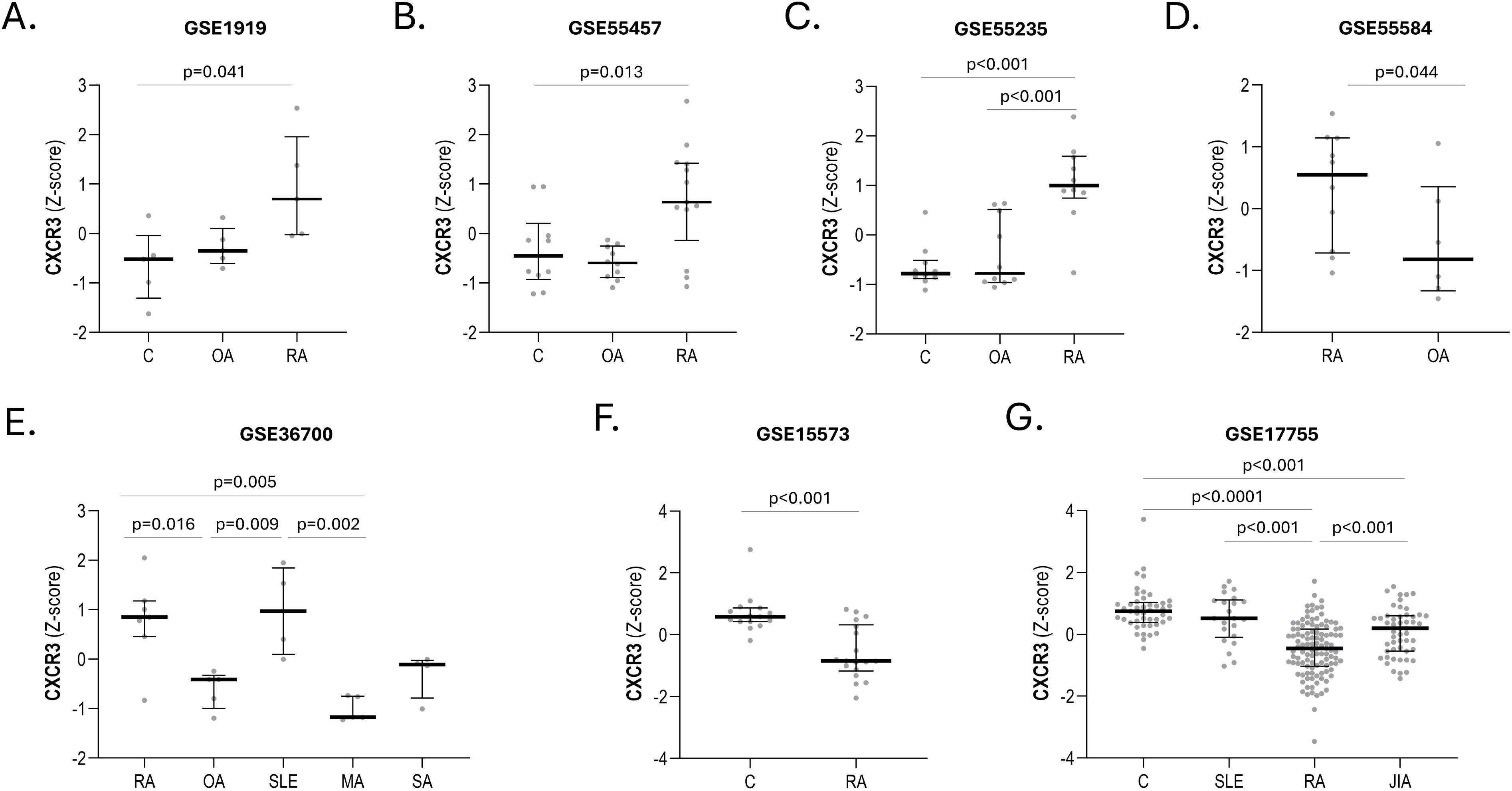
CXCR3 expression in systemic autoimmune diseases. CXCR3 expression was obtained from publicly available datasets from GEO. Datasets containing CXCR3 expression data from synovial biopsies (A-E) or peripheral blood (F-G) were analyzed. CXCR3 differential expression was confirmed by GEO2R (see Results section) within the whole dataset, and CXCR3 expression was extracted from each dataset, normalized (Z-score) and individually analyzed by conventional analyses (Kruskal-Wallis or Mann-Withney U tests, as appropriate). Bars represent 25th percentile (lower), median and 75th percentile (upper). P-values were indicated. Group labelling was maintained as per original nomenclature. C: control, JIA: juvenile idiopathic arthritis, MA: microcrystalline arthritis, OA: osteoarthritis, RA: rheumatoid arthritis, SA: seronegative arthritis.

Finally, CXCR3 expression in the systemic compartment was analyzed. GSE15573 revealed a differential CXCR3 gene expression (p(adj)=3.34·10-5) between RA (n=18) and controls (n=15) from peripheral blood mononuclear cell samples (Figure 4F). Similarly, results from GSE17755, containing peripheral blood samples from 112 RA patients, 22 SLE patients and 45 controls, found differences in CXCR3 expression (p(adj)=2.10·10-3) (Figure 4G). Subgroup analysis revealed diminished CXCR3 expression in RA compared to control group, and to a similar level than JIA patients (Figure 4G), hence confirming a specific CXCR3 downregulation in the systemic compartment in inflammatory arthritidies.

Collectively, analysis of multiple publicly available datasets consistently demonstrated compartment-specific alterations of CXCR3 expression in the setting of RA, namely increased CXCR3 expression at synovial tissue level, alongside a concomitant downregulation in the systemic compartment.

## DISCUSSION

Although conventionally regarded as aberrant immune responses primarily driving tissue damage, the conceptualization of autoantibodies has substantially evolved over recent decades. Herein we demonstrate that autoantibodies targeting CXCR3 were altered in systemic autoimmunity. Notably, reduced anti-CXCR3 were unrelated to disease duration and already detectable in the earliest stage of the disease. Anti-CXCR3 were linked with atherosclerosis burden and related to a variety of pathogenic proteomic pathways. To the best of our knowledge, this is the first study investigating regulatory autoantibodies in early arthritis and to demonstrate an association with atherosclerosis in autoimmunity.

Our findings consistently revealed decreased anti-CXCR3 autoantibody levels in RA and SjD, in line with prior evidence in SjD [24] and similar evidence suggested for other regulatory antibodies in giant-cell arteritis [25]. This observation points to a common immunoregulatory pathway involving the CXCR3 axis across autoimmune conditions, hence strengthening its biological relevance. This may align with the shared autoantibody signatures observed in neurological conditions [26]. Notably, the absence of an association with disease features, along with the findings from the CSA group, reinforce a potential role in disease initiation and triggering, and arguing against these being secondary to chronic inflammation or an epiphenomenon. This is in line with their role as natural components of the immune system, as recently proposed [27]. Altogether, these observations challenge the current view of autoantibodies in autoimmunity, and promote a shift from a purely pathogenic paradigm towards a “double-edge sword” framework, in which antibodies may also constitute an endogenous homeostatic system. The inverse association with poor therapeutic outcomes is in line with this interpretation, and mirrors similar results reported in ANCA-associated vasculitis [28]. Similarly, lower anti-CXCR3 antibodies were also predictors of lung function deterioration in SSc [9]. Although initially counterintuitive, observing decreased autoantibodies in autoimmunity is not unprecedented [29]. Additionally, a broad repertoire of IgG autoantibodies can be detected in the general population [30], with intrinsic properties of common autoantigens having been demonstrated, and contrasting with disease-related autoantibodies. Integration into antibody network landscapes is imperative to elucidate functional correlates of these findings. In this context, the concept of antibodyome emerges as a powerful tool to dissect mechanisms underlying altered levels of regulatory antibodies and to establish functional significance in autoimmunity.

A major breakthrough from our study was the robust association between anti-CXCR3 antibodies and atherosclerosis. Reduced anti-CXCR3 levels were consistently associated with atherosclerosis occurrence and extent both in RA as well as in SjD, hence demonstrating for the first time their involvement in the setting of autoimmunity. The validation of this result in a cohort of established RA patients with a history of CVD provides valuable insight in a two-fold manner. First, as it confirms this association with clinical overt CVD. Second, as it validates this association in a composite definition of CVD including non-ischaemic manifestations, thereby broadening the clinical spectrum of relevance. Mechanistic evidence linking CXCR3+ T-cells and cardiac remodelling further support this observation [31]. Although a potential role as diagnostic marker had been proposed [32,33], their contribution as clinical biomarker remained insufficiently explored. By conducting a comprehensive analysis of their incremental value, our data proved that anti-CXCR3 can be not only independent predictors but also yielded significant added value for risk stratification. Moreover, they can be combined with existing instruments for risk stratification, thereby addressing a major unmet need in the research agenda [34,35]. Finally, recent data from a large registry has unveiled a positive association between anti-CXCR3 antibodies and CV risk in the general population [8], which contrast with our findings. However, these discrepancies should be interpreted with caution, particularly in the light of the autoimmune background. Accumulating evidence has demonstrated that autoimmunity not only represents an additional risk factor itself (reviewed in [36]) but also substantially modifies the relative contribution of traditional CV risk factors [37,38]. Therefore, distinct mechanistic circuits operating may be key to reconcile these findings. Functional differences in antibodies between these two populations cannot be ruled out.

Although certain autoantibodies have been found to be decreased in atherosclerosis, mostly involved in clearance, the participation of regulatory targeting GPCRs calls for a novel framework. Experimental CXCR3 blockade or inactivation has been shown to dampen atherosclerosis progression [39], which is consistent with the protective effect of anti-CXCR3 herein suggested. CXCR3 is central to Th1-polarized lymphocyte migration towards sites of inflammation, and its ligands are strongly expressed in both inflamed synovium [40,41] and salivary glands [42], especially during disease exacerbation and flares. Therefore, the observed reduction in circulating anti-CXCR3 autoantibodies may rather reflect a selective accumulation within target tissues, thus leading to a loss of endogenous mechanisms that normally restrain chemokine receptor signalling and immune cell recruitment in the systemic compartment. An agonistic effect of anti-CXCR3 on immune cell migration in autoimmunity has been suggested [9]. Given that anti-CXCR3 antibodies may interfere with cell trafficking, decreased levels may in turn favour enhanced lymphocyte and monocyte transendothelial migration in the vasculature, thereby promoting atherosclerosis development. The consistent differential CXCR3 expression between synovial and systemic compartment in RA strongly supports this notion, also in line with the Th1 profile observed at the synovial level in RA [43]. This hypothesis may be also in line with the occurrence of shared mechanisms between joint inflammation and atherosclerosis development [44]. Of note, it may provide a functional insight into the connection between poor therapeutic outcomes and CV risk [45,46], also in accordance with our data. Of note, these findings provide robust observational evidence towards previous hypothesis [47]. Alternatively, low anti-CXCR3 antibodies might reflect enhanced binding to the endothelium. Endothelial cells upregulate CXCR3 upon inflammatory stimuli, especially in advance or unstable plaques [8], and anti-CXCR3 antibodies may therefore bind within the lesion and become sequestered or consumed. Preclinical models of atherosclerosis support this notion [48]. This ‘sink’ effect would in turn mean less systemic coverage of CXCR3+ cells before they encounter vascular chemokine gradients, thus enabling a more efficient CXCR3-dependent homing to sites of arterial inflammation. The associations with plaque characteristics in our study may support this hypothesis, which may be indicate higher influx of immune cells into plaques. Interestingly, increased CXCR3+ T-cells in atherosclerosis lesions have been described in lupus patients [49]. The combined enrichment of inflammatory, cell adhesion and chemotaxis pathways in our proteomic platform further reinforces this hypothesis. This hypothesis is also in line with the proposed role of anti-GPCRs antibodies as modulators of endothelial integrity and vascular-immune interface [5]. Additionally, regulatory autoantibodies can also exert their effects via receptor ligation and internalization [6], leading to tonic desensitization. Reduced anti-CXCR3 antibodies may consequently leave more CXCR3 available at the surface of immune cells, or may be insufficient to counterbalance high ligand production, thereby enhancing chemotactic responsiveness and transendothelial migration into plaques, favouring Th1 plaque accumulation and inflammation. A reduced competitive effect with ligand under low anti-CXCR3 conditions may also facilitate a more efficient binding of CXCR3 with endogenous chemokines to drive chemotaxis. Other mechanisms such as antibody consumption by complement activation may be ruled out, at least in part, according to our findings. This is consistent with the Fab-mediated mechanisms in predominating regulatory antibody functions compared to Fc-mediated predominance in their classical counterparts as well as with IgG subclass usage [6]. Finally, it is important to consider that antibody production may differ between patients and controls, so affinities and functions (agonist, antagonist or biased agonist) may also diverge, adding an additional layer of complexity. In sum, decreased anti-CXCR3 regulatory antibodies could plausibly favour enhanced CXCR3-dependent leukocyte recruitment into plaques, or altered receptor desensitization. These mechanisms, or a combination of thereof, may aggravate atherosclerosis progression in autoimmunity. Elucidating the exact functional phenotype of these antibodies and their compartmentalization, and passive transfer experiments in autoimmune and non-autoimmune models are warranted to provide experimental confirmation. Finally, the diversity of functional pathways retrieved for anti-CXCR3 underscores the central role of GPCRs signalling in driving disease mechanisms and highlight the contribution of non-traditional CV risk mechanisms. Delineating individual GPCRs signatures is imperative to explain individual susceptibility and encourages integrative analysis to decipher functional networks.

The characterization of anti-CXCR3 antibodies in relation to clinical outcomes in RA and SjD paves the way for a comprehensive investigation of anti-GPCRs antibodies in the broader context of autoimmunity. Given the central role of GPCRs in regulating a plethora of biological processes, these antibodies may shed new light into systemic manifestations, such as neurological outcomes including fatigue, dysautonomia or cognitive dysfunction [5], which represent critical determinants of quality of life. Furthermore, deciphering regulatory antibody networks in these conditions will likely inform complex, multi-system interactions underlying autoimmune pathogenesis at the systemic level [7]. Recent data from other clinical scenarios, including limited evidence from SjD, are reassuring in this regard [24]. Whether disturbed antibody networks may represent a viable therapeutic target in autoimmunity is yet to be determined. Although our findings failed to exhibit significant changes in antibody levels upon TNFi exposure, it is conceivable that these fluctuations inform changes in antibody networks that may be of biological and clinical relevance, as reported in other complex systems [50].

In conclusion, altered anti-CXCR3 antibodies may represent a common hallmark of autoimmunity. Disturbed anti-CXCR3 levels could yield new insight into disease pathogenesis, clinical implications and broad antibody biology in rheumatic diseases. Clinical translation of anti-CXCR3 antibodies hold promise to improve risk stratification beyond traditional scores. Collectively, these findings align with the emerging concept that components of the regulatory autoantibody repertoire can exert immunomodulatory functions and that network-wide disruptions may contribute to immune dysregulation and clinical outcomes in rheumatic autoimmune diseases. This study comes with certain limitations such as cross-sectional design or lack of functional data. Validation of CXCR3 expression came from independent cohorts, so future paired studies are awaited. Prospective trials to assess added value in hard clinical endoints (i.e. MACE) are warranted. This study opens new avenues towards system-level approaches integrating multi-layered characterization of regulatory antibody networks to improve our understanding of systemic autoimmunity and identifying potential therapeutic targets. A reframing of the current concept of autoantibodies in autoimmunity within this expanded, regulatory framework is encouraged.

## Author contributions

All authors were involved in drafting the manuscript or revising it critically for important intellectual content and all the authors gave their approval of the final version of the manuscript to be published.

Study conception and design: JRC, GR, AS

Acquisition of data: DM-P, MAL, AIPA, SSD, SAC, AS, JRC

Analysis and interpretation of data: DM-P, AS, GR, JRC

## Data Availability Statement

The data underlying this article are available in the article and in its online supplementary material.

## Acknowledgements

The authors would like to thank the “Liga Reumatológica Asturiana” for their collaboration and support.

