## Supplementary Material for "Reduced circulating anti-CXCR3 antibodies as a common hallmark bridging systemic autoimmunity and atherosclerosis"

**SUPPLEMENTARY MATERIALS**

Supplementary Material and Methods

Supplementary Tables

Supplementary Figures

**SUPPLEMENTARY TABLES**

**Supplementary Table 1: Demographic and clinical features of study participants.** The demographic and clinical features of the participants entered in this study were summarized. Variables were expressed as median (interquartile range) or n(%), unless otherwise stated, according to the distribution of the variables.

|  | **HC**  **n=65** | **CSA n=14** | **RA  n=82** |
| --- | --- | --- | --- |
| Age (years), mean (range) | 52.12  (36.10-71.20) | 49.28  (30.00-62.33) | 58.51  (30.33-83.67) |
| Gender (female/male) | 50/15 | 14/0 | 66/16 |
| ***Clinical features*** |  |  |  |
| Duration of symptoms (weeks) |  | 24.00 (40.00) | 20.00 (22.00) |
| Morning stiffness (minutes) |  | 30.00 (50.00) | 60.00 (80.0) |
| Tender joint count |  | 3.00 (3.00) | 8.00 (7.00) |
| Swollen joint count |  | 0.00 (1.00) | 6.00 (5.00) |
| Patient global assessment (VAS 0-100) |  | 30.00 (50.00) | 70.00 (25.00) |
| DAS28 |  |  | 5.40 (1.78) |
| DAS28 at 6 months (n=58) |  |  | 2.60 (1.94) |
| DAS28 at 12 months (n=58) |  |  | 2.19 (2.07) |
| HAQ |  | 0.55 (0.60) | 1.11 (1.00) |
| Pain (VAS 0-10) |  | 5.00 (5.00) | 7.00 (2.00) |
| RF+, n(%) |  | 8 (66.6) | 57 (69.5) |
| ACPA+, n(%) |  | 7 (58.3) | 56 (68.2) |
| ANA+, n(%) |  | 2 (14.2) | 34 (41.4) |
| ***Laboratory parameters*** |  |  |  |
| ESR (mm/h) |  | 7.50 (8.84) | 24.00 (27.00) |
| CRP (mg/dl) |  | 0.15 (0.30) | 0.80 (2.20) |
| Total cholesterol (mg/dl) |  | 196.50 (44.00) | 193.00 (48.00) |
| HDL-cholesterol (mg/dl) |  | 74.00 (15.59) | 50.00 (19.00) |
| LDL-cholesterol (mg/dl) |  | 116.00 (49.00) | 115.20 (47.00) |
| Triglycerides (mg/dl) |  | 74.50 (54.00) | 107.50 (80.00) |
| IgG (g/l) |  | 10.03 (3.33) | 10.99 (3.28) |
| IgA (g/l) |  | 1.97 (1.41) | 2.70 (1.38) |
| IgM (g/l) |  | 1.11 (0.73) | 1.11 (0.61) |
| C3 (g/l) |  | 1.23 (0.34) | 1.33 (0.42) |
| C4 (g/l) |  | 0.22 (0.08) | 0.29 (0.12) |
| ***Traditional CV risk factors, n(%)*** |  |  |  |
| Hypertension |  | 1 (12.5) | 28 (34.1) |
| Diabetes |  | 0 (0.0) | 9 (10.9) |
| Dyslipidemia |  | 3 (37.5) | 24 (29.2) |
| Smoking |  | 10 (71.4) | 31 (27.8) |
| Obesity (BMI>30 kg/m^2^) |  | 3 (37.5) | 33 (40.2) |
| ***Atherosclerosis burden*** |  | n=13 | n=77 |
| Plaque presence, n(%) |  | 4 (30.7) | 46 (59.7) |
| Plaque number, mean (range) |  | 0.46 (0-3) | 0.96 (0-4) |
| High-risk plaques, n(%) |  | 0 (0.0) | 20 (25.9) |
| cIMT (mm) |  | 0.58±0.15 | 0.67±0.10 |

**Supplementary Table 2: Demographic and parameters of established RA patients.** The demographic and clinical features of established RA patients recruited and matched HC as a validation cohort were shown. Variables were expressed as median (interquartile range), mean±SD or n(%), unless otherwise stated, according to the distribution of the variables.

|  | **HC**  **n=80** | **Established RA  n=103** |
| --- | --- | --- |
| Age (years), mean (range) | 51.05  (21.00 – 69.00) | 55.12  (22.00-87.00) |
| Gender (female/male) | 65/15 | 84/19 |
| ***Clinical features*** |  |  |
| Disease duration, median (range) (years) |  | 4.12 (0.50 – 21.13) |
| Age at diagnosis (years), mean (range) |  | 40.85 (18.00 – 80.92) |
| Tender joint count |  | 2.00 (6.50) |
| Swollen joint count |  | 1.00 (4.00) |
| Patient global assessment (VAS 0-100) |  | 30.00 (50.50) |
| Pain (VAS 0-10) |  | 3.00 (2.12) |
| DAS28 |  | 3.15 (2.25) |
| HAQ |  | 0.87 (1.14) |
| RF+, n(%) |  | 55 (53.3) |
| ACPA+, n(%) |  | 66 (64.0) |
| ANA+, n(%) |  | 46 (44.6) |
| History of CV events, n(%) |  | 16 (17.7) |
| ***Laboratory parameters*** |  |  |
| ESR (mm/h) |  | 15.00 (25.00) |
| CRP (mg/dl) |  | 1.00 (4.00) |
| Total cholesterol (mg/dl) |  | 203.00 (55.00) |
| HDL-cholesterol (mg/dl) |  | 62.00 (18.75) |
| LDL-cholesterol (mg/dl) |  | 118.00 (46.00) |
| Triglycerides (mg/dl) |  | 104.00 (73.25) |
| ***Traditional CV risk factors*** |  |  |
| Hypertension, n(%) |  | 38 (36.8) |
| Diabetes, n(%) |  | 8 (7.7) |
| Dyslipidemia, n(%) |  | 31 (30.1) |
| Smoking, n(%) |  | 30 (29.1) |
| Obesity (BMI>30 kg/m^2^), n(%) |  | 18 (17.4) |
| ***Treatments, n(%)*** |  |  |
| Glucocorticoids |  | 63 (61.1) |
| Methotrexate |  | 75 (72.8) |
| TNF blockers |  | 42 (40.7) |
| IL-6 blockers |  | 9 (8.7) |
| Leflunomide |  | 11 (10.6) |

**Supplementary Table 3: Demographic and parameters of SjD patients.** The demographic and clinical features of SjD patients recruited and matched HC were shown. Variables were expressed as median (interquartile range), mean±SD or n(%), unless otherwise stated, according to the distribution of the variables.

|  | **HC**  **n=45** | **SjD  n=44** |
| --- | --- | --- |
| Age (years), mean (range) | 63.51  (25.30-79.00) | 61.36  (35.00-80.00) |
| Gender (female/male) | 40/5 | 40/4 |
| ***Clinical features*** |  |  |
| Disease duration, median (range) (years) |  | 8.12 (5.10 – 25.01) |
| Xerostomia |  | 43 (97.7) |
| Xeroftalmia |  | 44 (100.0) |
| Extra-glandular manifestations |  | 31 (70.4) |
| ESSDAI |  | 5.11±5.08 |
| clinESSDAI |  | 4.45±4.74 |
| ANA+, n(%) |  | 39 (88.6) |
| Anti-RO/SSA+, n(%) |  | 38 (86.4) |
| Anti-LA/SSB+, n(%) |  | 16 (36.4) |
| RF+, n(%) |  | 25 (56.8) |
| ***Laboratory parameters*** |  |  |
| ESR (mm/h) |  | 20.00 (28.00) |
| CRP (mg/dl) |  | 1.00 (1.15) |
| Total cholesterol (mg/dl) |  | 100.50 (69.80) |
| HDL-cholesterol (mg/dl) |  | 60.50 (19.50) |
| LDL-cholesterol (mg/dl) |  | 117.00 (55.50) |
| Triglycerides (mg/dl) |  | 82.50 (60.00) |
| IgG (g/l) |  | 14.30 (8.02) |
| IgA (g/l) |  | 2.31 (1.31) |
| IgM (g/l) |  | 1.10 (0.74) |
| C3 (g/l) |  | 1.13 (0.38) |
| C4 (g/l) |  | 0.20 (0.14) |
| ***Traditional CV risk factors*** |  |  |
| Hypertension, n(%) |  | 13 (29.5) |
| Diabetes, n(%) |  | 5 (11.4) |
| Dyslipidemia, n(%) |  | 6 (13.6) |
| Smoking, n(%) |  | 8 (18.2) |
| Obesity (BMI>30 kg/m^2^), n(%) |  | 10 (22.7) |
| ***Treatments, n(%)*** |  |  |
| HCQ |  | 23 (52.3) |
| Glucocorticoids |  | 14 (31.8) |
| Methotrexate |  | 4 (9.1) |
| Azathioprine |  | 3 (6.8) |
| Rituximab |  | 3 (6.8) |
| ***Atherosclerosis burden*** |  |  |
| Plaque presence, n(%) |  | 22 (50.0) |
| Plaque number, mean (range) |  | 0.67 (0-4) |
| cIMT (mm) |  | 0.62±0.10 |

**Supplementary Table 4: Associations between anti-CXCR3 levels and cytokines.** Associations between serum levels of anti-CXCR3 antibodies and cytokines in early RA were analysed by Spearman’s rank tests in early RA. Coefficients (r) and p-values are shown. Those reaching statistical significance were highlighted in bold.

|  | **Anti-CXCR3** |
| --- | --- |
| IFNa | r=-0.055  p=0.622 |
| IL-6 | r=-0.097  p=0.388 |
| TNF | r=-0.162  p=0.145 |
| IFNg | r=-0.147  p=0.187 |
| IL-1b | r=-0.068  p=0.545 |
| IL-23 | r=-0.092  p=0.413 |
| IL-12 | r=-0.123  p=0.269 |
| IL-33 | r=-0.079  p=0.478 |
| IL-10 | r=-0.082  p=0.461 |
| IL-17 | r=0.169  p=0.130 |
| IL-8 | r=-0.008  p=0.942 |
| IL-18 | r=0.024  p=0.828 |
| IL-21 | r=-0.080  p=0.550 |
| APRIL | r=-0.046  p=0.733 |
| BAFF | r=-0.061  p=0.648 |

**Supplementary Table 5: Demographic and clinical features of RA patients undergoing TNFi treatment.** The demographic, clinical features and treatment outcomes at 3 months of biological-naïve RA patients undergoing TNFi were summarized. Variables were expressed as median (interquartile range), mean±SD or n(%), unless otherwise stated.

|  | **Biological-naïve RA n=13** | |
| --- | --- | --- |
| Age (years), mean (range) | 46.71  (30.75-65.42) | |
| Gender (female/male) | 12/1 | |
| ***Clinical features*** |  |  |
| Disease duration, median (range) (years) | 1.54 (1.00 – 7.17) | |
| Age at diagnosis (years), mean (range) | 44.28 (29.08 – 62.75) | |
| RF+, n(%) | 5 (38.4) | |
| ACPA+, n(%) | 6 (46.1) | |
| ANA+, n(%) | 5 (38.4) | |
| ***Treatment outcomes*** | *Baseline* | *3 months* |
| Tender joint count | 9.00 (5.00) | 4.50 (3.00) |
| Swollen joint count | 5.00 (3.00) | 3.00 (1.69) |
| ESR (mm/h) | 13.00 (28.00) | 9.5 (16.25) |
| CRP (mg/dl) | 3.00 (4.15) | 1.00 (1.50) |
| Patient global assessment (VAS 0-100) | 66.00 (17.00) | 35.00 (20.28) |
| Pain (VAS 0-10) | 6.00 (2.90) | 3.25 (2.97) |
| DAS28 | 5.15 (1.99) | 3.80 (1.97) |
| HAQ | 1.20 (0.79) | 0.87 (0.85) |

**Supplementary Table 6: Anti-CXCR3 antibodies as predictors of atherosclerosis in SjD.** The role of anti-CXCR3 levels as predictor of atherosclerosis occurrence in SjD patients was analysed by univariate and multivariate logistic regression models. Sex was omitted from the multivariate analysis due to sample size limitations. Odds ratio (OR), 95% confidence intervals (95% CI) and p-values for each predictor were shown. Those reaching statistical significance were highlighted in bold.

|  | **OR** | **95% CI** | **p-value** |
| --- | --- | --- | --- |
| ***Univariate*** | | | |
| Anti-CXCR3, per unit | 0.826 | 0.684 – 0.996 | **0.046** |
| ***Multivariate*** | | | |
| Anti-CXCR3, per unit | 0.748 | 0.567 – 0.988 | **0.041** |
| Age, per year | 1.297 | 1.090 – 1.545 | **0.003** |
| Smoking, yes | 0.739 | 0.088 – 6.215 | 0.739 |
| Hypertension, yes | 0.761 | 0.117 – 4.797 | 0.750 |
| Diabetes, yes | 0.607 | 0.004 – 84.140 | 0.843 |
| Dyslipidemia, yes | 1.290 | 0.010 – 17.728 | 0.919 |

**SUPPLEMENTARY FIGURE LEGENDS**

**Supplementary Figure 1: Proteomic analyses of anti-CXCR3 autoantibodies in SjD, atherosclerosis status and disease overlap.** Analysis of proteomic hits associated with anti-CXCR3 antibodies in SjD were performed. Protein-protein interaction networks (A) and protein and process enrichment analysis (B) were conducted as in the RA group (Figure 3). SjD and RA proteomic outputs were analyzed by heatmap (C) and network term layout (D). Colour code represents RA (red) and SjD (blue). Within SjD, group-wise comparisons among SjD, SjD patients with atherosclerosis, and SjD atherosclerosis-free patients were performed in heatmap (E) and network term layout (F) as conducted in the RA group (Figure 3). Colour code represents whole SjD group (red) and SjD patients with atherosclerosis (blue).
